# Holistic Structural Visualization of Coronary Arteries with Single-catheter Dual-modality Intravascular Imaging Combining IVUS and OFDI

**DOI:** 10.1101/2020.06.19.20130302

**Authors:** Jian Ren, Milen Shishkov, Martin Villiger, Kenichiro Otsuka, Seemantini Nadkarni, Brett Bouma

## Abstract

**Objectives:** To test the imaging performance and practical use of a novel dual-modality intravascular imaging system combining intravascular ultrasound (IVUS) and optical coherence tomography (OCT) into a single catheter.

**Background:** IVUS enables assessing coronary plaque burden, a robust metric for patient prognosis, while OCT and OFDI provide high-resolution images of coronary microstructure and detailed assessment of stent implantation. Owing to their complementary strengths, co-registering IVUS and OFDI provides a more comprehensive assessment of coronary lesions during PCI.

**Methods:** We developed a 2.6Fr imaging catheter integrating both IVUS and OFDI, interfacing to a dual-modality imaging console through a fast interchange connector. A novel algorithm fuses the IVUS and OFDI signals into a single combined image. We verified the performance of the two modalities and their visualization by imaging of cadaveric coronary arteries and tested practical imaging in the catheterization laboratory in swine *in vivo*.

**Results:** Coronary atherosclerotic lesions of cadaver hearts revealed complementary optical and acoustic image features. Spatial co-registration of the modalities was confirmed by high correlation of measured lumen areas. Fused into a combined visualization, dual-modality imaging offers quantitative characterization of lesions, including plaque-burden and fibrous cap thickness. Imaging *in vivo* does not add procedural complexity compared to conventional single modality imaging.

**Conclusion:** Dual-modality IVUS/OFDI imaging with fused visualization provides improved assessment of coronary atherosclerotic lesions and is compatible with a routine clinical setting. Combining the strength of the two modalities offers unique opportunities both as a powerful research instrument and to improve clinical management of patients undergoing PCI.

**Visual abstract:** 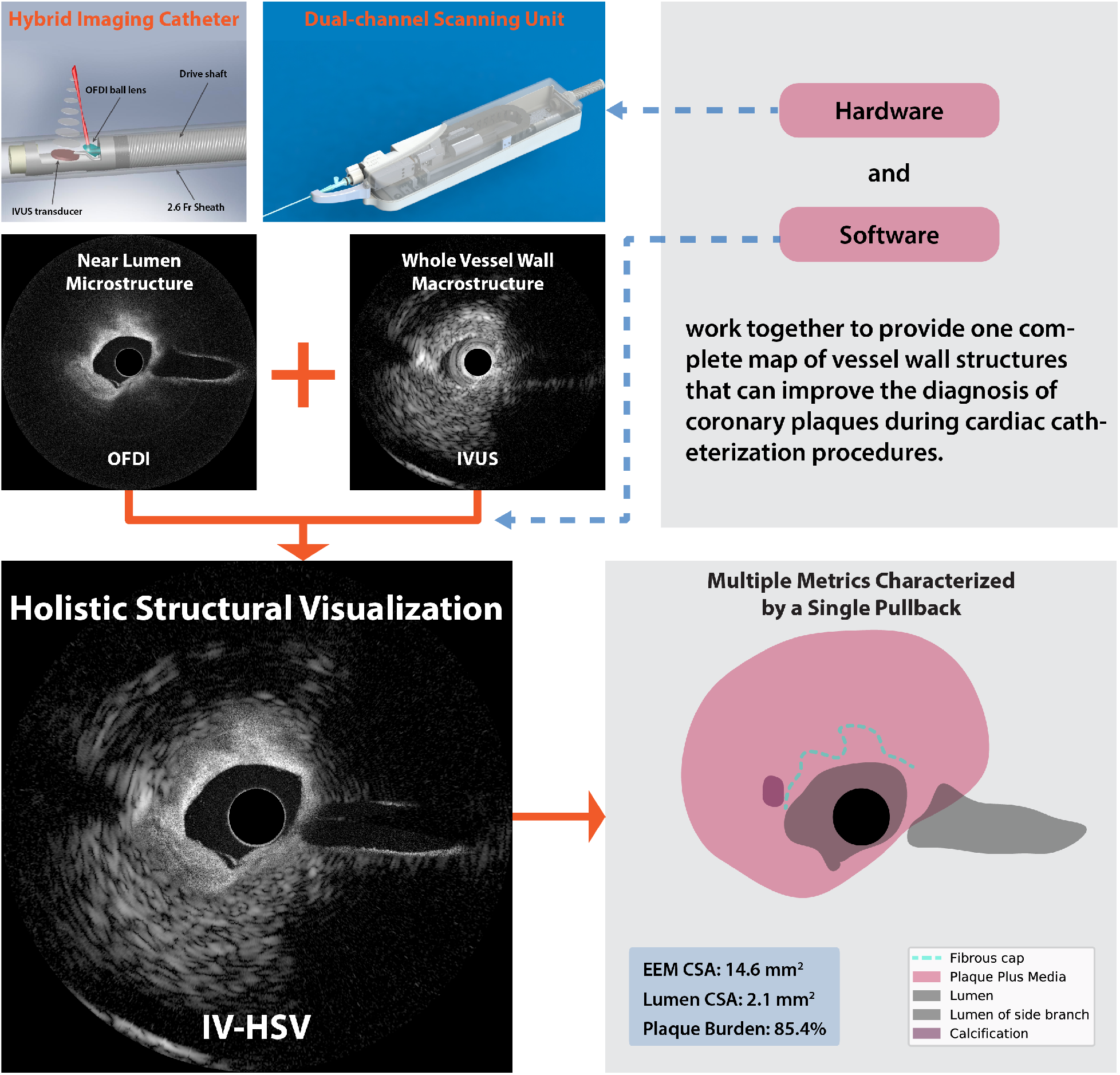

**Highlights:**

▪ A catheter-based imaging system integrating ultrasound and optical frequency domain imaging enables real time intravascular holistic structural visualization (IV-HSV) of coronary arterial lesions with a single pullback during cardiac catheterization procedures.
▪ A computational method to automatically fuse images from both modalities generates a single complete map of vessel wall structures - the IV-HSV image, which offers holistic investigation of coronary plaque key features, such as fibrous cap thickness, calcification, and plaque burden.
▪ The visualization of plaque morphology along the entire extent of the coronary arterial wall reveals vital information for guiding percutaneous coronary interventions and for advancing our understanding of the pathophysiology of coronary artery diseases.

**Condensed abstract:** To facilitate a holistic investigation of coronary arterial lesions for both basic research and clinical interventions, this work presents a catheter-based imaging system integrating IVUS and OFDI, as well as a new rendering method that computationally fuses the intrinsically co-registered images from both modalities into a single cross-sectional map of vessel structures. Imaging human cadaveric coronary arteries shows the benefit of this system by revealing near-lumen microstructures and spatially correlated macrostructures deep inside the vessel wall. The co-registration accuracy and operation in a clinical setting of this system was demonstrated through swine catheterization *in vivo*.

## Introduction

The role of intravascular ultrasound (IVUS) and optical coherence tomography (OCT) in the guidance of percutaneous coronary intervention (PCI) in patients with coronary artery disease has been the subject of extensive investigation (1–4). These intravascular imaging modalities have also deepened our understanding of the pathophysiology of acute coronary syndromes (ACS) and vascular tissue response to stent implantation and drug therapy (5–7). The deep signal penetration of IVUS affords imaging through even the most stenotic lesions and enables measuring coronary plaque burden, which has been shown to be a robust predictor of long-term cardiovascular outcomes (8,9). OCT modalities, such as optical frequency domain imaging (OFDI), reveal near lumen microstructure at a much higher resolution of 10-15 µm (10,11), detecting fine structural features of coronary atherosclerosis and implanted stents, that are clinically relevant to identify the culprit lesion and guide stent sizing (3). Yet, no single modality provides complete visualization of coronary plaque, including delineation of fibrous cap, identification of calcification, and evaluation of lesion depth (12).

Post mortem and clinical studies using both IVUS and OCT/OFDI have demonstrated that their combination improves detection of thin-capped fibroatheromas (TCFAs) (13,14) and enables accurate biomechanical modelling of plaque structural stress (15–17). However, the use of independent, standalone IVUS and OCT/OFDI instruments employed in these studies is impractical in a routine clinical setting, entails potentially imperfect co-registration, and impairs real-time analysis. To address these limitations, dual-modality imaging systems have been recently developed to conduct simultaneous IVUS and OCT/OFDI imaging through a single hybrid imaging catheter. From the proof-of-concept benchtop demonstration (18) to the first-in-human images (19), a number of developments have been achieved to increase the frame rate (20), improve the image quality, and miniaturize the catheters of varying distal designs (21– 25). Tested in a wide range of vascular specimens, these combined systems have confirmed their potential for clinical adoption (19,26).

We developed an integrated IVUS-OFDI system that implements critical technical improvements to enable simple and reliable operation in cardiac catheterization laboratories. Our system consists of an imaging console offering concurrent OFDI and IVUS video rate imaging with a pullback speed of up to 20 mm/s. The clinical grade catheters comprise an optical ball lens and an ultrasonic transducer in a 2.6 Fr, low-profile design. A dual-channel rotary joint provides parallel optical and electrical connections and allows for a fast interchange of the disposable catheters. Furthermore, we developed a computational method to automatically fuse the co-registered IVUS-OFDI cross-sectional images, providing macro- and micro-structural features of coronary atherosclerotic plaques in a single image. Here, we present the dual-modality IVUS-OFDI system and report on imaging experiments with human cadaver hearts and swine *in vivo*, demonstrating the unique benefits of this hybrid imaging system in the catheterization laboratory.

## Methods

### Integrated IVUS-OFDI imaging system

#### Dual-modality catheter

To ensure compatibility with PCI procedures in clinical practice, the hybrid catheter was engineered to integrate the imaging components for both modalities into a single narrow-gauge assembly. As illustrated in Figure 1A, the distal end incorporates an optical ball-lens with a diameter of ∼200 µm and a single-element ultrasound transducer with a side dimension of 0.5 mm. The angle-polished surface of the ball-lens is metalized to deflect the optical beam towards the vessel wall at a focal distance of 2.6 mm from the catheter center. The optical and ultrasound beams are aligned by adjusting the orientations of the transducer and the angle-polished surface during catheter assembly. The two imaging components are encased in an open metal housing with an overall outer diameter of 0.6 mm and a rigid length of 1.3 mm. A single mode optical fiber guides OFDI light between the ball-lens and a self-aligning angle-polished fiber connector at the proximal end of the catheter. The ultrasonic transducer is electrically connected to a customized radio-frequency (RF) connector at the proximal end via a pair of 34-gauge copper wires. The optical fiber and the RF wires are bonded to a torque cable that serves as the drive shaft for the circumferential scanning. The entire assembly fits into a clinical-grade sheath, which has a 2.6 Fr/3.2 Fr 132 cm monorail design compatible with standard 0.014” guidewires (Figure 1B). The proximal end connectors are disclosed in the inset of Figure 1B.

**Figure 1.**
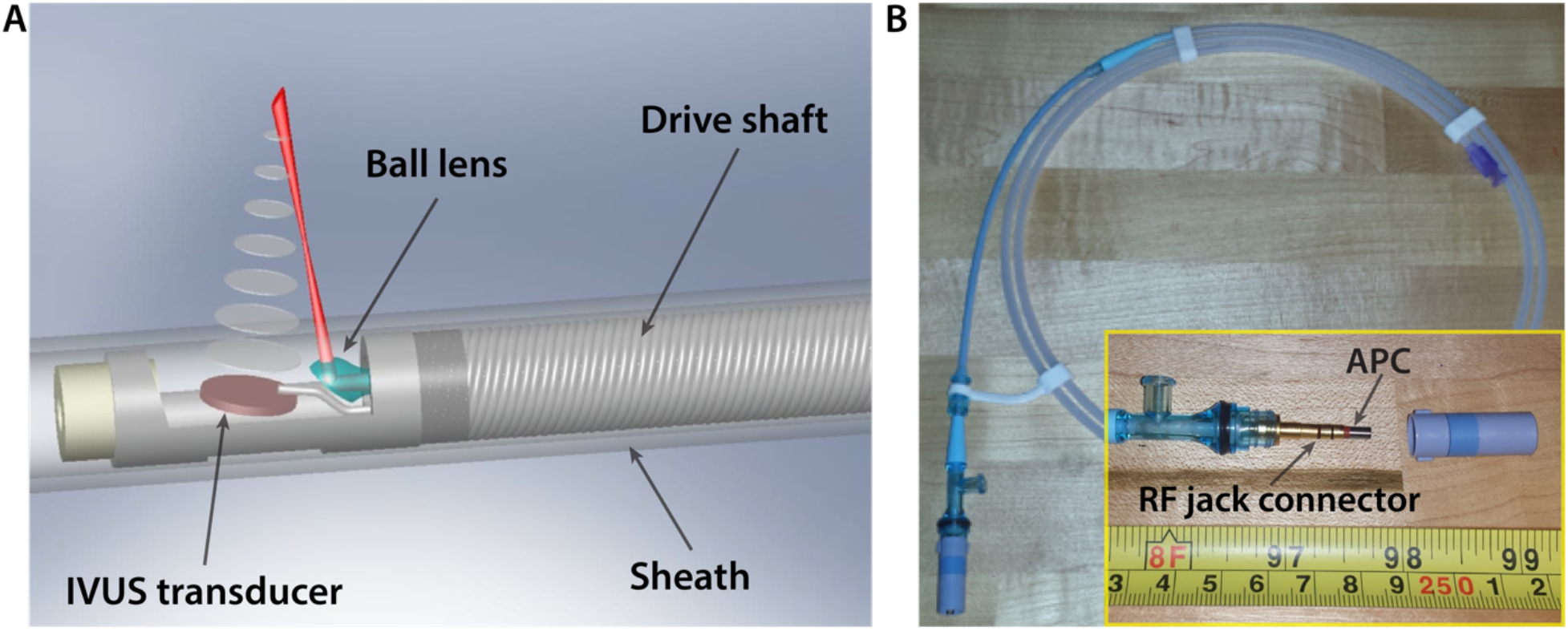
Dual-modality catheter integrating IVUS and OFDI. **(A)** Schematic rendering of the distal end. **(B)** Photograph of an assembled and packaged clinical-grade catheter. The inset displays the RF electrical connection and the angled physical contact (APC) fiber connector.

#### Dual-channel scanning unit

A hybrid optical-electrical rotary interface was developed to connect the catheter comprising the spinning dual-modality imaging probe to the static imaging console for concurrent transmission of both OFDI and IVUS signals. A mechanical locking mechanism is employed to secure the dual-channel connection with a simple click-on, facilitating fast interchange of the disposable catheters between catheterization procedures. Two motor drive axes drive the hybrid rotary interface. One of them is used to spin the hybrid rotary interface supporting >100 rotations per second. The rotary interface and its driving motor are mounted on a customized linear stage. The second motor translates the stage and the connected imaging probe relative to the static outer sheath to achieve pullbacks with a travel range of 20 cm and a speed ranging from 5-20mm/s. To enhance the system reliability, a motor control subsystem operates independently of the imaging console dedicated to signal acquisition and processing. It coordinates all motion sequences, including catheter rotation, pullback, automatic advancing for resetting catheter position, and optimal positioning for catheter interchange.

#### Dual-modality imaging console

The console integrates the OFDI and IVUS subsystems into a single mobile cart. The OFDI subsystem utilizes a custom-made wavelength-swept laser, providing axial scans at 50 kHz repetition rate and delivering 10 mW of optical power onto the tissue. The 110 nm wavelength range of this source is centered at 1300 nm for optimal cardiac tissue penetration, offering an axial resolution of ∼7 μm. Balanced and polarization-diverse detection of the OFDI interference signal avoids polarization fading and yields a system sensitivity of 106 dB. The IVUS subsystem employs a customized pulser/receiver operating at a carrier frequency of 40 MHz with a fractional bandwidth of 50 %. The pulser/receiver operates at a repetition rate of 25 kHz derived from the 50 kHz OFDI trigger signal using a standalone programmable input/output device. This timing mechanism results in the synchronized acquisitions of both modalities at a ratio of 2:1 (OFDI/IVUS). Adjusting the catheter rotation to acquire 1024 IVUS axial scans and 2048 OFDI axial scans per frame results in a frame rate of 24.8 frames per second. A custom-developed C++ application continuously streams the digitized data to a RAID hard-drive array, ensuring uninterrupted data acquisition. A subset of the data is processed and visualized in real-time, achieving processing and rendering of about 22 frames per second. While the real-time performance varies slightly depending on the instantaneous CPU load, the data streaming and recording have highest priority to avoid data loss and ensure offline analysis of the complete dataset. The application also includes routines and graphical user-interfaces to configure and control the various system components, and communicates with the control subsystem of the scanning unit.

### Intravascular holistic structural visualization (IV-HSV)

We developed a computational method to fuse inherently co-registered IVUS-OFDI image pairs into single structural images of the vessel wall, representing subsurface microstructure with OFDI and transitioning to IVUS for depths beyond the reach of the OFDI signal (Figure 2). First, the OFDI and IVUS cross-sectional images are scaled to represent actual dimensions by considering the refractive index and the acoustic impedance of the flushing medium. After this geometric calibration, the border where the OFDI signal starts to transition to the IVUS signal is automatically determined by analyzing the OFDI intensity radial profile. An intensity threshold and a thickness parameter are used to locate the depth where the OFDI signal is depleted or is most likely dominated by multiply scattered light. The error function with a width of 90μm is used to define a smooth transition, by creating weight masks that are applied to the OFDI and IVUS images. The final fused image is the weighted sum of the image pair using these masks.

**Figure 2.**
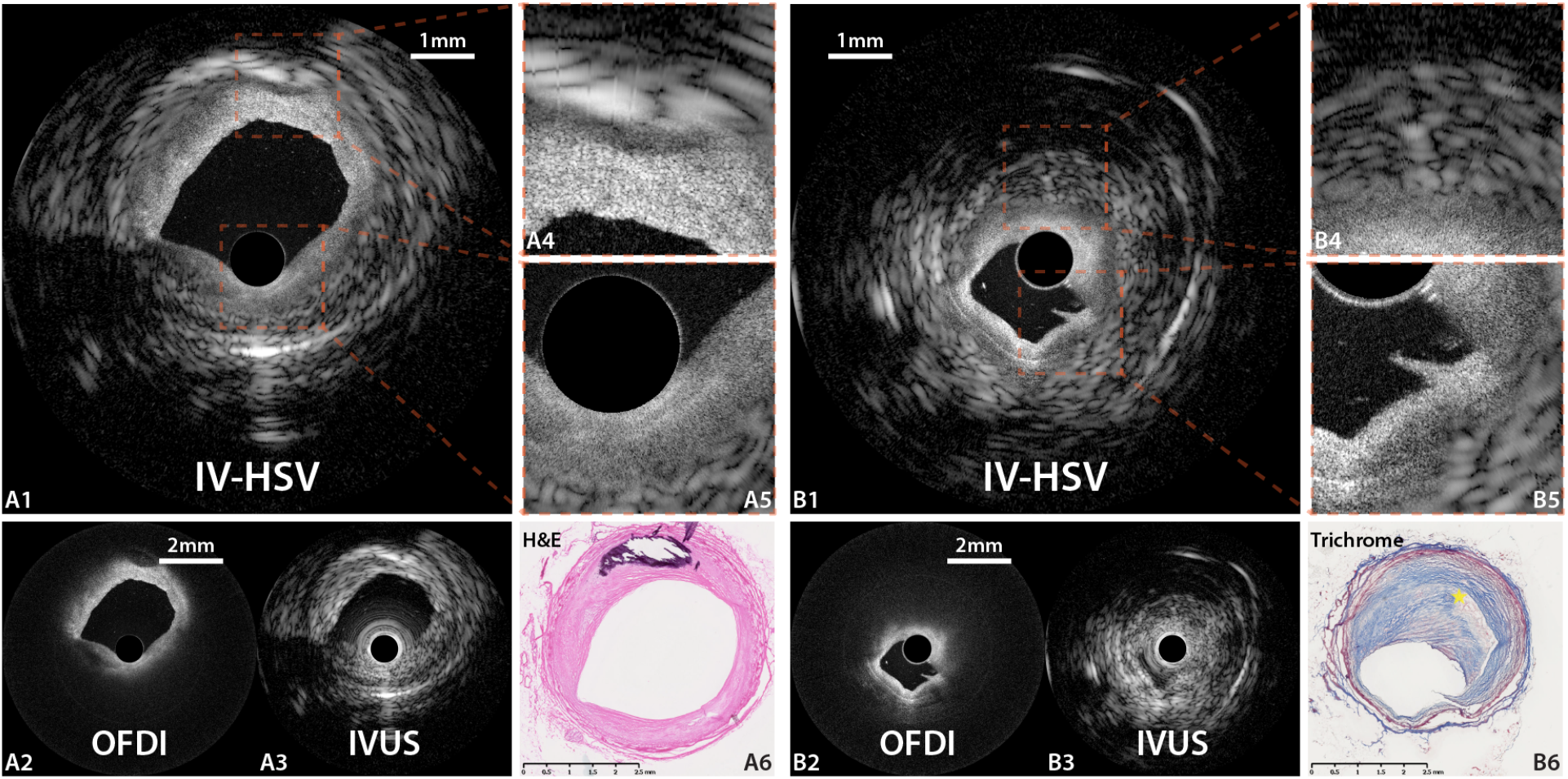
Holistic structural visualization (IV-HSV) of coronary plaques *ex vivo*. Representative images of a fibro-calcified plaque **(A1 to A6)** and a necrotic core fibroatheroma **(B1 to B6)**. IV-HSV images **(A1 and B1)** generated by automatically fusing the OFDI **(A2 and B2)** and IVUS **(A3 and B3)** cross-sectional images. **(A4 and A5)** and **(B4 and B5)** show the magnified views of the square areas of interest indicated in **(A1)** and **(B1)**, respectively. **(A6 and B6)** Matching histopathology, stained with hematoxylin and eosin (**A6)**, and trichrome **(B6)**, respectively. Scale bars in **(A2, B2)** apply to both **(A2 and A3)** and **(B2 and B3)**, respectively.

### Intracoronary imaging in cadaveric human heart

Cadaver human hearts were accrued from the National Disease Research Interchange under a protocol approved by the institutional review board at the Massachusetts General Hospital. Hearts were collected within 24 hours after death and stored at -80 degree Celsius. Tissues were properly thawed before imaging experiments. Left anterior descending artery (LAD), left circumflex artery (LCX), and right coronary artery segments with perivascular tissues were resected from the hearts. A custom fixture was used to mount the artery segments and ensure accurate co-registration with histopathology (27). The proximal end of the segments was connected to a Y-shape Luer connector, allowing for both saline or contrast flushing and catheter access. Side branches were ligated to avoid fluid leakage. The catheter was deployed with a 0.014-inch guidewire that was withdrawn for imaging. During pullback imaging, saline was manually flushed through the segments.

After imaging, frozen sections of the arterial samples were prepared for histopathology. Arterial segments were snap frozen within their fixtures. To facilitate accurate co-registration, the fixtures contain vertical slots to enable cutting of the tissue blocks into smaller segments. Two consecutive 7 µm-thick sections were collected every 1 mm along the artery segments and processed for hematoxylin-eosin (H&E) and Trichrome staining, respectively. Each slide was digitized with NanoZoomer (Hamamatsu Photonics, K.K.) for further analysis.

### Live swine catheterization and imaging

For practical testing of the dual-modality system in the cardiac catheterization lab at the Massachusetts General Hospital, we conducted intravascular imaging *in vivo* in a healthy Yorkshire pig under deep anesthesia. Vital signals were monitored throughout the entire procedure. A 7 Fr guide catheter was placed through the carotid artery to gain access to the coronary arteries. Both the LAD and LCX were catheterized using a 0.014-inch guidewire (Figure 5A). A total of fourteen 60 mm-long pullbacks were acquired at various pullback speeds (5-20 mm/s). Contrast injection was manually performed to displace circulating blood in the imaged coronary artery to ensure good image quality for OFDI. The study followed the regulations of Massachusetts General Hospital and with generally accepted guidelines governing such work.

### IV-HSV image analysis

The fused dual-modality images visualize multiple important anatomic structures that are relevant for characterizing atherosclerotic lesions (visual abstract). We segmented the lumen, calcifications, fibrous cap boundary, and side-branches in OFDI images and the EEM in IVUS images. Based on the segmentations, the percent plaque plus media CSA (%Plaque burden) was calculated as:

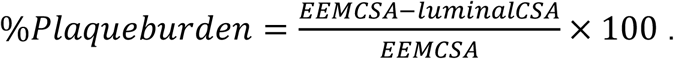

The fusion processing, the image rendering, and the calculation of the CSAs and the plaque burden were all automated in Python.

To demonstrate the accuracy of the intrinsic co-registration of this dual-modality imaging system, we compared the luminal CSA measured independently in the properly scaled OFDI and IVUS cross-sectional images using Pearson’s correlation analysis.

The lumen was manually segmented with custom-built MATLAB routines. Evaluation of the areas and statistical analysis were performed with custom Python routines and the SciPy.stat package.

## Results

### Visualization of coronary plaques with dual-modality imaging

First, we imaged a human cadaver heart with the dual-modality system, where various coronary plaque features were visualized. Demonstrating typical fibroatheromata, Figure 3 displays the IV-HSV images, generated by optimally merging the intrinsically co-registered OFDI and IVUS cross-sectional images, together with the segmentation of individual plaque features and matching histology. The IV-HSV image (Figure 3-A1) clearly renders the lumen shape and the fibrous cap as captured by OFDI, complemented by deeper plaque structures and the entire circumference of the vessel wall disclosed by IVUS. Notably, the fusion algorithm automatically selected OFDI to depict the near-lumen microstructure of the vessel injury at 6-8 o’clock with high resolution, which is more challenging to identify in the IVUS image. Furthermore, segmentation of the IV-HSV image enabled quantitative measurements of luminal CSA and EEM CSA providing the plaque burden of the fibroatheroma as percentage (Figure 3-A4 and B4). The cross-section in Figure 3B is located close to a side branch. While both OFDI and IVUS cross-sectional images visualize the side branch at 3 o’clock - an important anatomical landmark during PCI - the OFDI image reveals its morphology with higher fidelity. Determined by the fusion algorithm, the side branch is represented by its OFDI appearance in the IV-HSV image, preferable for providing the best assessment of vessel morphology.

**Figure 3.**
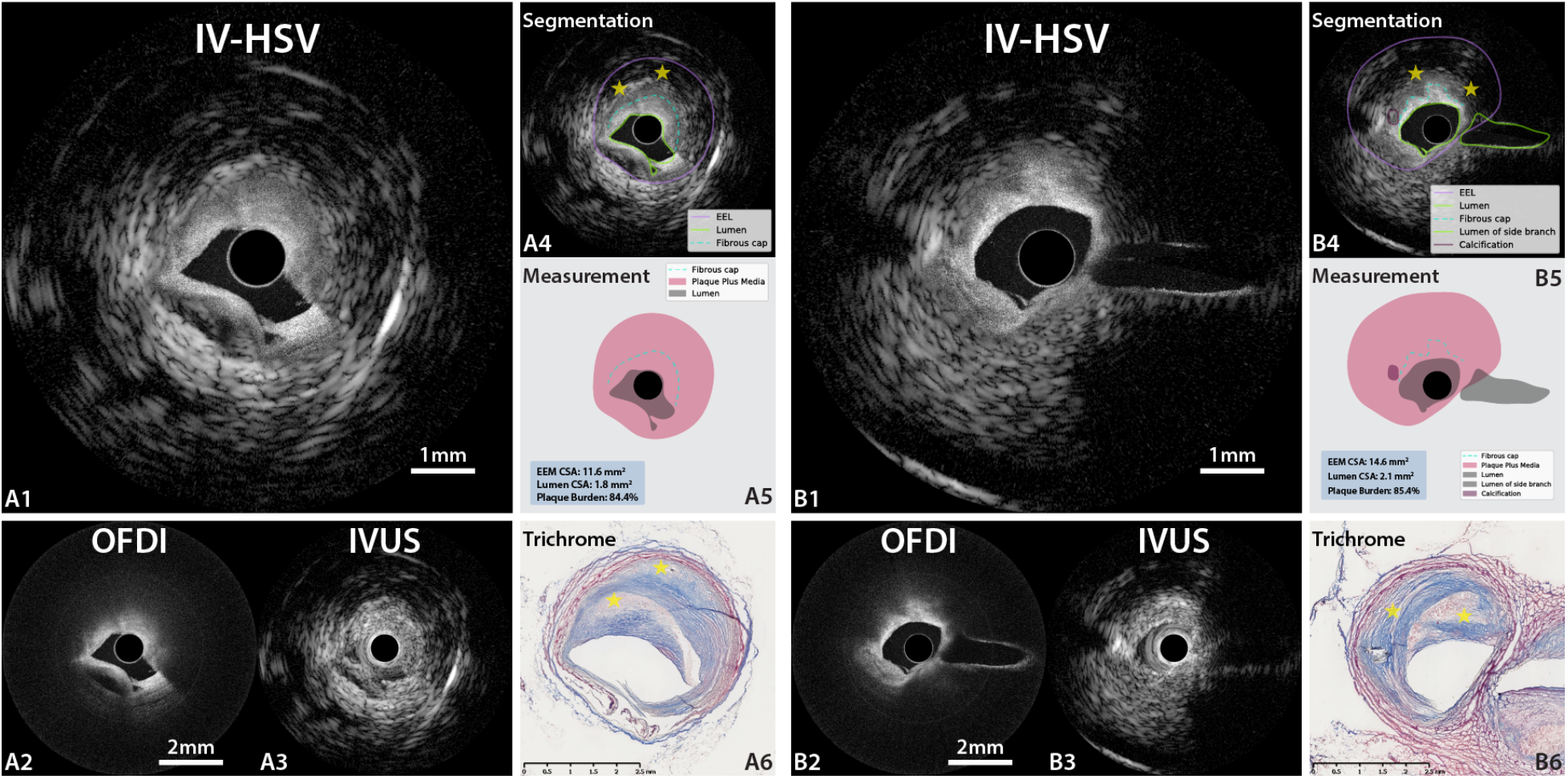
Representative holistic structural visualization (IV-HSV) of fibroatheromas in human coronary artery *ex vivo*. IV-HSV images **(A1 and B1)**, combining the OFDI images **(A2 and B2)** and IVUS images **(A3 and B3). (A4 and B4)** Segmentation of various plaque features in the IV-HSV images, and isolated visualization of the segmented structures, enabling computation of plaque burden **(A5 and B5). (A6 and B6)** Matching trichrome-stained histopathology. **A2 to A5** share the scale bar in **A2**, and **B2 to B5** share the scale bar in **B2. (A1-A5)** A dissection of the coronary artery appears from 6 to 8 o’clock. **(A1)** The IV-HSV image visualizes all relevant details of the dissected vessel wall by optimally merging the IVUS and OFDI appearances. **(B1)** The IV-HSV image clearly visualizes the lumen and the entire vessel wall, including a side branch and calcification. The yellow stars in **A4** and **B4** indicate necrotic cores, identified by the IVUS signal and confirmed by histopathology (yellow stars in **A6** and **B6**, respectively).

Figure 4A displays a further interesting example of an atherosclerotic lesion. The conventional OFDI image suggests a lipid-rich plaque and discloses a thrombus-like structure on the luminal surface (Figure 4-A2). Histopathology confirmed the presence of newer intimal tissue on top of a pathological intimal thickening (Figure 4-A6). While the OFDI appearance mimicked a fibroatheroma, the co-registered IVUS image clearly depicts the EEM behind a lesion with only a modest amount of lipid. However, the newer intimal structure clearly visualized in the OFDI image is challenging to appreciate in the IVUS image (Figure 4-A3). Owing to the complementarity of the underlying imaging modalities, the IV-HSV image reveals structures across the entire diameter of the vessel wall and with high resolution close to the lumen, enabling the accurate estimation of plaque burden (Figure 4-A4 and -A5).

**Figure 4.**
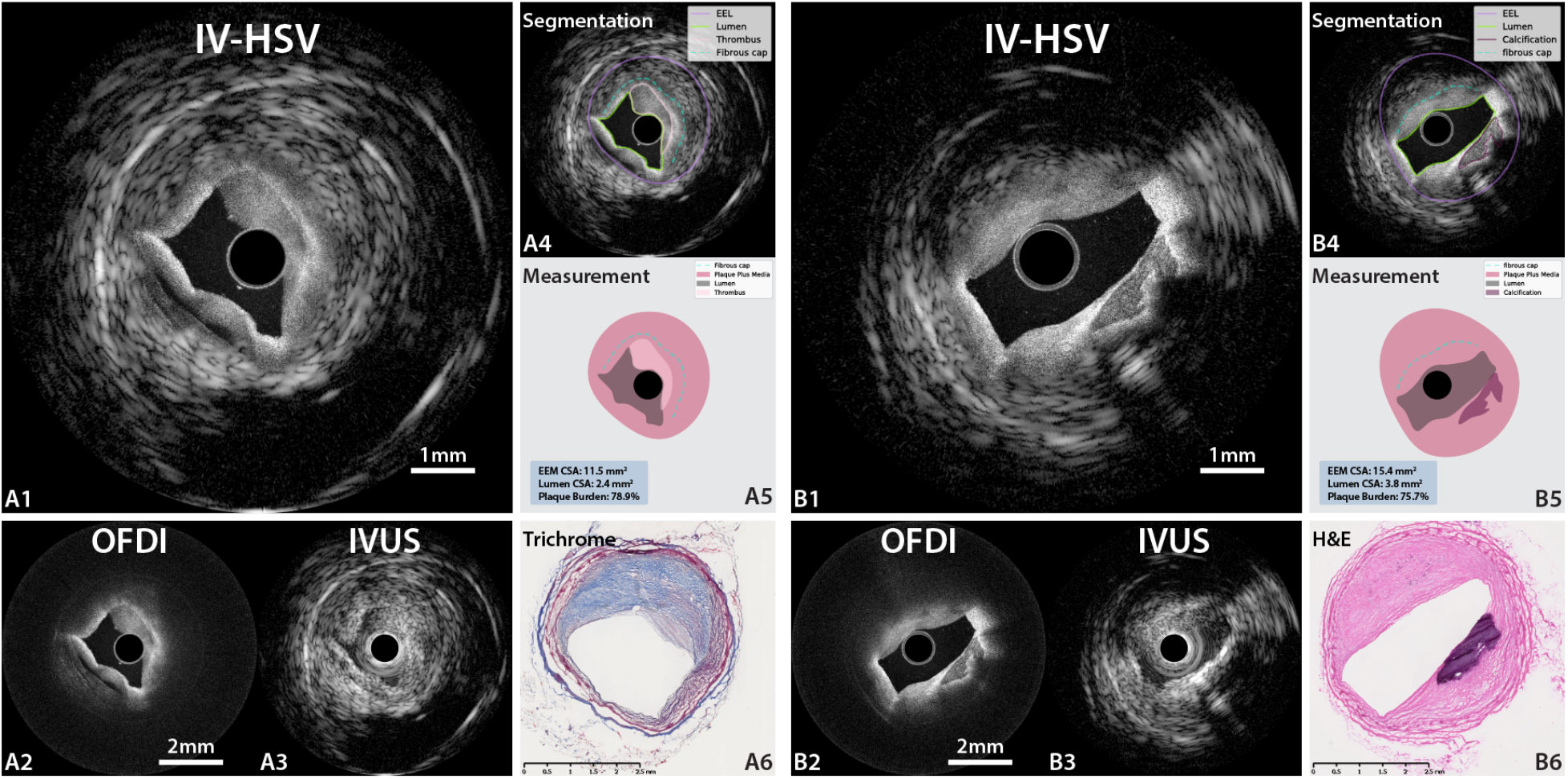
Representative holistic structural visualization (IV-HSV) of a pathological intimal thickening and a fibro-calcified plaque in human coronary artery ex vivo. IV-HSV images **(A1 and B1)**, combining the OFDI images **(A2 and B2)** and IVUS images **(A3 and B3). (A4 and B4)** Segmentation of various plaque features in the IV-HSV images, and isolated visualization of the segmented structures, enabling computation of plaque burden **(A5 and B5)**. Matching histopathology, stained with trichrome (**A6)**, and hematoxylin and eosin **(B6)**, respectively. **(A1)** The IV-HSV image visualizes the subsurface microstructures and the EEM. **(B1)** The IV-HSV image reveals the shape of the superficial calcification and the necrotic core as a region with echo-attenuation.

Figure 4B presents a fibro-calcified plaque with lipid pool. The OFDI image (Figure 4-B2) delineates the detailed geometry of the calcification and the upper boundary of the lipid pool, in close agreement with histology (Figure 4-B6). In OFDI, calcifications appear translucent and with a well-defined border. In comparison, the large difference in acoustic impedance between calcified and surrounding tissue casts an acoustic shadow that prevents IVUS from imaging behind calcified tissue or determining its depth (Figure 4-B3). Combined, the IV-HSV image (Figure 4-B1) clearly visualizes the arterial lumen and the EEM of the vessel, facilitating the characterization of various plaque features within a single map.

### Dual-modality IVUS-OFDI in swine coronary artery in vivo

Next, we examined the utility and performance of the dual-modality IVUS-OFDI system in the cardiac catheterization laboratory. Figure 5 provides an overview of the imaging with the system in the coronary arteries of a healthy swine. Figure 5D, 5E, 5G, and 5H show intrinsically co-registered dual-modality IVUS-OFDI cross-sectional images acquired in the LAD, at a pullback rate of 10 mm/s. Both OFDI (Figure 5D and 5G) and IVUS (Figure 5E and 5H) images show matching side branch locations. In addition, the Pearson’s correlation analysis (Figure 5B) demonstrates an excellent correlation between luminal CSA measured with OFDI and IVUS (r = 0.97, p < 0.001). The Bland-Altman plot (Figure 5C) suggests neither the measurement bias (-0.07mm^2^) nor the limits of agreement (0.25 and -0.38mm^2^) is clinically important. Both Figure 5B and 5C validated the intrinsic co-registration of the two modalities.

**Figure 5.**
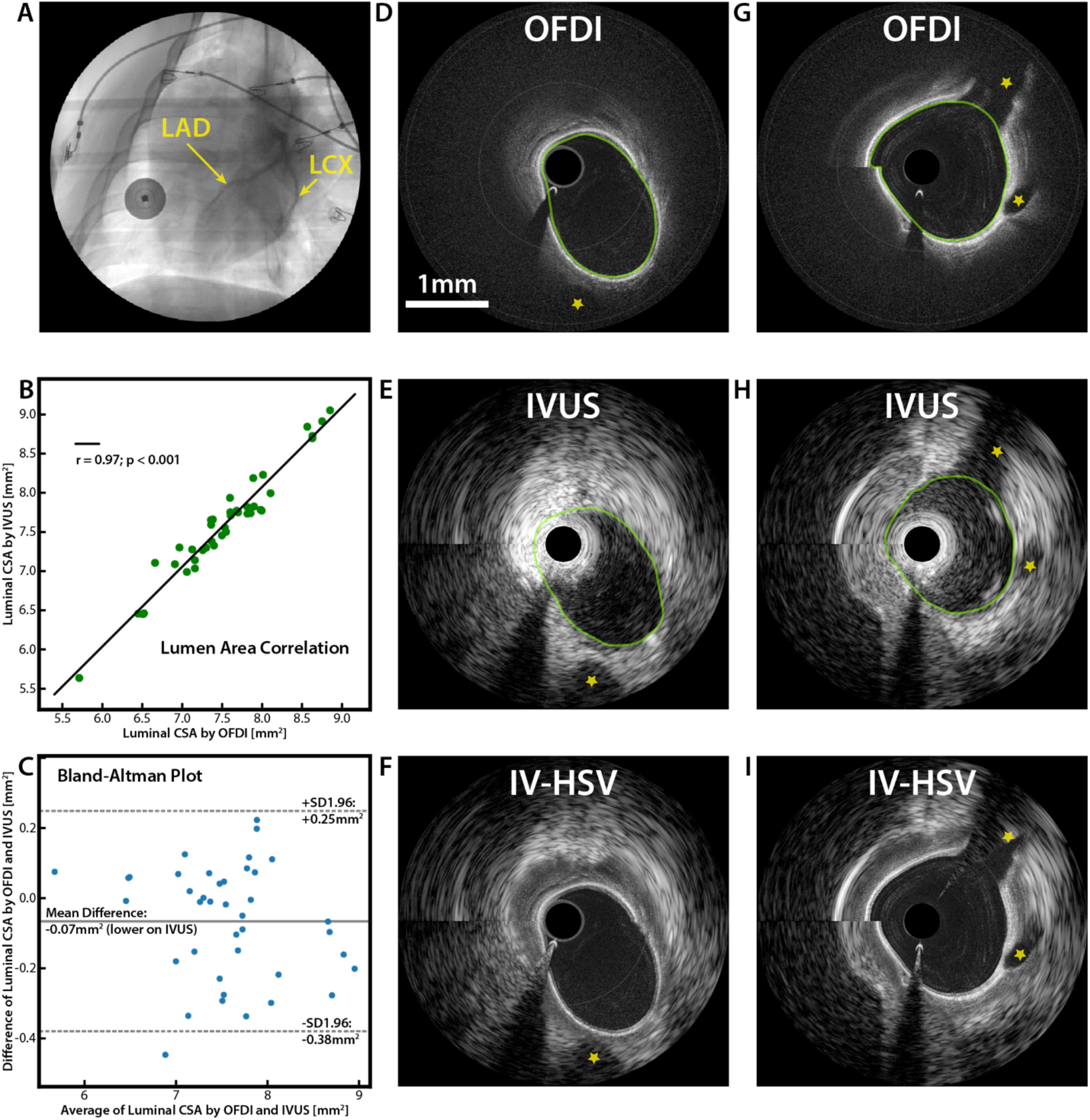
*In vivo* swine catheterization and intravascular imaging. **(A)** Coronary angiogram showing the left coronary arteries during pullback imaging. **(B)** Pearson’s correlation analysis of luminal CSA measured independently with OFDI and IVUS. **(C)** Bland-Altman plot of the luminal CSA measured independently with OFDI and IVUS. **(D, E, and F)** and **(G, H, and I)** are intrinsically co-registered OFDI, IVUS, and IV-HSV cross-sectional images acquired in the LAD. The yellow stars indicate the locations of side branches. Both **(B)** and **(C)** show excellent agreement between measurements with the two modalities, accounting to the accuracy of the intrinsic co-registration of the dual-modality imaging system. The green lines in **(D, E, G, and H)** are the lumen segmentations. **(D-I)** share the same scale bar in **(B)**.

## Discussion

The present study reports on the development of an intravascular dual-modality imaging system integrating IVUS and OFDI into a single catheter enabling fast interchange required for ease of operation in the catheterization laboratory. We further developed a novel computational method to fuse the cross-sectional images from the two modalities for convenient visualization of the macro- and microstructures of coronary plaques – IV-HSV. Imaging of swine coronary arteries *in vivo* with the dual-modality system demonstrated accurate intrinsic co-registration of cross-sectional OFDI and IVUS images acquired simultaneously with a single pullback, which is unattainable by using the two imaging modalities sequentially. Translated to the clinic, the IV-HSV of this dual-modality IVUS-OFDI system would provide a holistic assessment of coronary arterial lesions and may help refine intervention and management strategies for patients undergoing PCI.

### Development of the dual-modality system

The physical alignment of the optical and acoustic beams ensures spatial co-registration of the IVUS and OFDI axial scans. The absence of rotational or longitudinal offsets between the two modalities found in previous work (rotational 90° (25), 180° (24), longitudinal 2mm (23)), eliminates potential rotational co-registration errors due to non-uniform rotational distortion as well as longitudinal co-registration errors due to cardiac motion. Furthermore, the synchronization scheme of the acquisition system guarantees temporal co-registration of the axial scans of both modalities. Both the temporal synchronization and the physical beam alignment contribute to the highly accurate correlation of vessel structures in the obtained images. It is worth noting that there are small longitudinal and angular offsets between the optical and acoustic beams, as shown in Figure 1A. The angular offset is introduced to compensate the longitudinal offset so that two beams have the largest overlap along the imaging depth. Compared to IVUS resolution, the overall misalignment is sufficiently small to be ignored, as confirmed by Figure 5B and 5C.

The integration of both optical and acoustic sensing into a single catheter required special considerations for its distal design. Ultrasound transmission requires immersion of the distal assembly in an echolucent medium, requiring flushing of the open-ended catheter sheath with saline. Therefore, the optical components inevitably need to operate in the same aqueous environment. In contrast to conventional OFDI catheters, where refraction happens at the glass-air interface of the ball lens within a sealed catheter sheath, the refractive index (RI) surrounding the ball lens is increased in the present case from 1 (air) to about 1.3 (water), a value much closer to the RI of the silica-based ball lens (1.43). This posed two challenges. First, conventional OFDI catheters employ total internal reflection at the angle-polished surface of the ball lens to deflect the beam towards the vessel wall. This effect is frustrated when immersed in a higher RI and ceases to take place. Instead, we metalized the angle-polished surface to cause the beam to deflect regardless of the surrounding mediums’ RI. Second, the reduced RI contrast between the medium and the glass lowers the refractive power of the curved interface. To maintain a high-quality near-lumen optical focus, we significantly reduced the diameter of the ball lens to about 200 µm. Although this dimension approaches its lower limit – the diameter of the fiber (125 µm), our fabrication process produced ball lenses with very consistent quality.

### IV-HSV of coronary plaques and vessels

Dual-modality IVUS-OFDI imaging offers unique opportunities for both research and clinical use by characterizing both micro- and macro-structures of coronary atherosclerotic lesions with a single imaging procedure. OFDI has been utilized to resolve various intravascular structures at a resolution of <20 µm. Nevertheless, its imaging depth is limited to 1-2 mm by tissue scattering. On the other hand, IVUS penetrates through the entire vessel wall, offering estimation of plaque burden and insight into the composition of large plaques. However, its inferior resolution of several hundred microns prevents the visualization of fine tissue microstructures. In light of the individual modalities’ complementary traits, the proposed IV-HSV imaging provides a more comprehensive examination of coronary atherosclerotic lesions. Supplemental table 1 compares the capability of IVUS, OFDI and the dual-modality system of visualizing key features of the coronary arterial wall and implanted stents.

A first example demonstrating the unique benefits of IV-HSV is the estimation of plaque burden. Plaque burden measured with IVUS has been shown to be a robust predictor of short- and long-term cardiovascular outcomes in patients with coronary artery disease (8,9,28). Accurate estimation of plaque burden with OCT/OFDI in lipid-rich lesions remains challenging due to insufficient imaging penetration. Efforts using computational algorithms to estimate plaque burden from OCT/OFDI data has shown limited success (29–31). The dual-modality system offers a potentially more accurate estimation of plaque burden than either IVUS or OCT/OFDI alone, because it combines the robust mapping of the EEM available to IVUS with the accurate delineation of the lumen owing to the improved resolution of OCT/OFDI.

Dual-modality imaging may also improve the detection of rupture-prone plaques. Despite significant efforts in advancing intravascular imaging modalities, accurate identification of these plaques in patients remains difficult. Clinical studies have indicated that a large number of TCFAs diagnosed by OCT/OFDI do not cause acute coronary events (32,33). While OCT/OFDI excels at identifying thin fibrous caps, it is prone to overestimating lesion severity owing to its limited imaging depth and it can mis-classify pathological intimal thickening as fibroatheroma. The combined use of IVUS and OFDI has been shown to improve the accuracy of identifying TCFAs (13,14). The automatically generated IV-HSV images may facilitate real-time detection of rupture-prone TCFAs with increased accuracy and offer improved risk assessment of no-reflow phenomenon and clinically relevant periprocedural myocardial infarction during PCI.

While previous technical developments enabled the demonstration of the dual modal concept and its First-in-Human application, the current work embodies critical technical improvements and innovations, including: 1) The new distal probe design offers improved sensor alignment, which, together with the precise timing of image acquisition, results in accurately co-registered IVUS/OFDI image pairs. The refined fabrication processes of sensor elements and the comprehensive optimization of the OFDI and IVUS subsystems contribute to the high image quality. In addition, the innovations in the hybrid rotary joint and catheter packaging ensure consistent system performance for practical application environments. 2) The novel IV-HSV processing synergistically integrates structural maps from both modalities, providing the clinical-relevant information in real-time. Besides the basic processing and control functionalities, as offered by previous systems, the software systems of the imaging console and the stand-alone motor control unit were customized to maximize safety and usability. Combined, these advances resulted in an intravascular visualization tool with unprecedented image quality and compatible with routine clinical use.

### Study limitations

First, imaging of coronary atherosclerosis was performed in a single of cadaver heart. Intracoronary imaging *in vivo* was performed in a healthy swine without coronary artery disease. Imaging in patients will be needed to demonstrate robust dual-modality imaging in a clinical setting. Second, the present study did not include imaging of stented coronary arteries. Further studies are needed to investigate whether the fusion algorithm is able to conveniently render stent struts and in-stent neointima for improved investigation of the mechanisms underlying stent thrombosis and in-stent restenosis. Third, the current fusion algorithm entirely relies on the OFDI signal to define the transition from OFDI to IVUS appearance. While this strategy works well for the majority of coronary lesions, it could result in non-optimal partitioning of the two modalities due to artifacts in OFDI images. Utilizing the joint information from both modalities, potentially coupled with prior pathophysiological knowledge in the form of a trained deep learning model holds promise to yield more robust and clinical-relevant visualization.

Finally, similar to conventional intravascular OCT/OFDI systems, the dual-modality system requires blood clearing through the injection of contrast media or saline for OFDI imaging. Operation without flushing would preclude OFDI image interpretation, but offer the ability to survey vessels using IVUS alone to locate lesions of interest for targeted dual-modality imaging. This would reduce the amount of used contrast medium and limit the risk of contrast-induced acute kidney injury during PCI (34).

## Conclusions

This study demonstrates the feasibility and prospects of dual-modality IVUS and OCT imaging using an integrated imaging system for the characterization of coronary arterial atherosclerotic lesions. Use of a single clinical-grade, dual-modality imaging catheter and real-time IV-HSV ensure convenient operation in a clinical setting. Owing to the synergy between IVUS and OFDI, IV-HSV imaging offers unique opportunities to study the etiology of coronary atherosclerosis and assess the risk of coronary lesions and clinical outcomes of patients undergoing PCI.

## PERSPECTIVES

### COMPETENCY IN MEDICAL KNOWLEDE

IVUS and OFDI are well-established intravascular imaging methods for guiding PCI. IVUS visualizes the entire vessel wall also in areas of disease, while OFDI images near-lumen microstructures of coronary arteries at high resolution. Combining insight from independent imaging with IVUS and OFDI has been shown to improve the detection of high-risk plaques and the estimation of plaque structural stress, but is impractical in a routine clinical setting. Here, we present a catheter-based imaging system integrating IVUS and OFDI enabling real-time IV-HSV of coronary arteries with a single catheter. IV-HSV images represent automatically fused IVUS and OFDI images in a single complete map of macro- and microstructures of the coronary arterial wall.

### TRANSLATIONAL OUTLOOK

The utility of the dual-modality IVUS-OFDI imaging system should be tested in patients with coronary artery disease undergoing PCI.

## Data Availability

The data that support the findings of this study are available from the corresponding author upon reasonable request

## Acknowledgements

Kenichiro Otsuka acknowledges partial support from the Japan Heart Foundation / Bayer Yakuhin Research Grant Abroad, the Uehara Memorial Foundation Postdoctoral Fellowship, and the Japan Society for the Promotion of Science Overseas Research Fellowship. The authors acknowledge Leon Ptaszek MD, PhD and Moussa Mansour, MD at the Department of Cardiology, Massachusetts General Hospital for *in vivo* swine imaging; Diane Tshikudi, MS and Pallavi Doradla, PhD at the Wellman Center for Photomedicine, Massachusetts General Hospital for their assistance in cadaver tissue preparation and histology.

## Abbreviations and Acronyms

ACS: acute coronary syndrome
CSA: cross sectional area
EEM: external elastic membrane
IV-HSV: intravascular holistic structural visualization
IVUS: intravascular ultrasound
LAD: left anterior descending artery
LCX: left circumflex artery
OCT: optical coherence tomography
OFDI: optical frequency domain imaging
PCI: percutaneous coronary intervention
TCFA: thin-capped fibroatheroma

---

**Supplementary Table 1.**
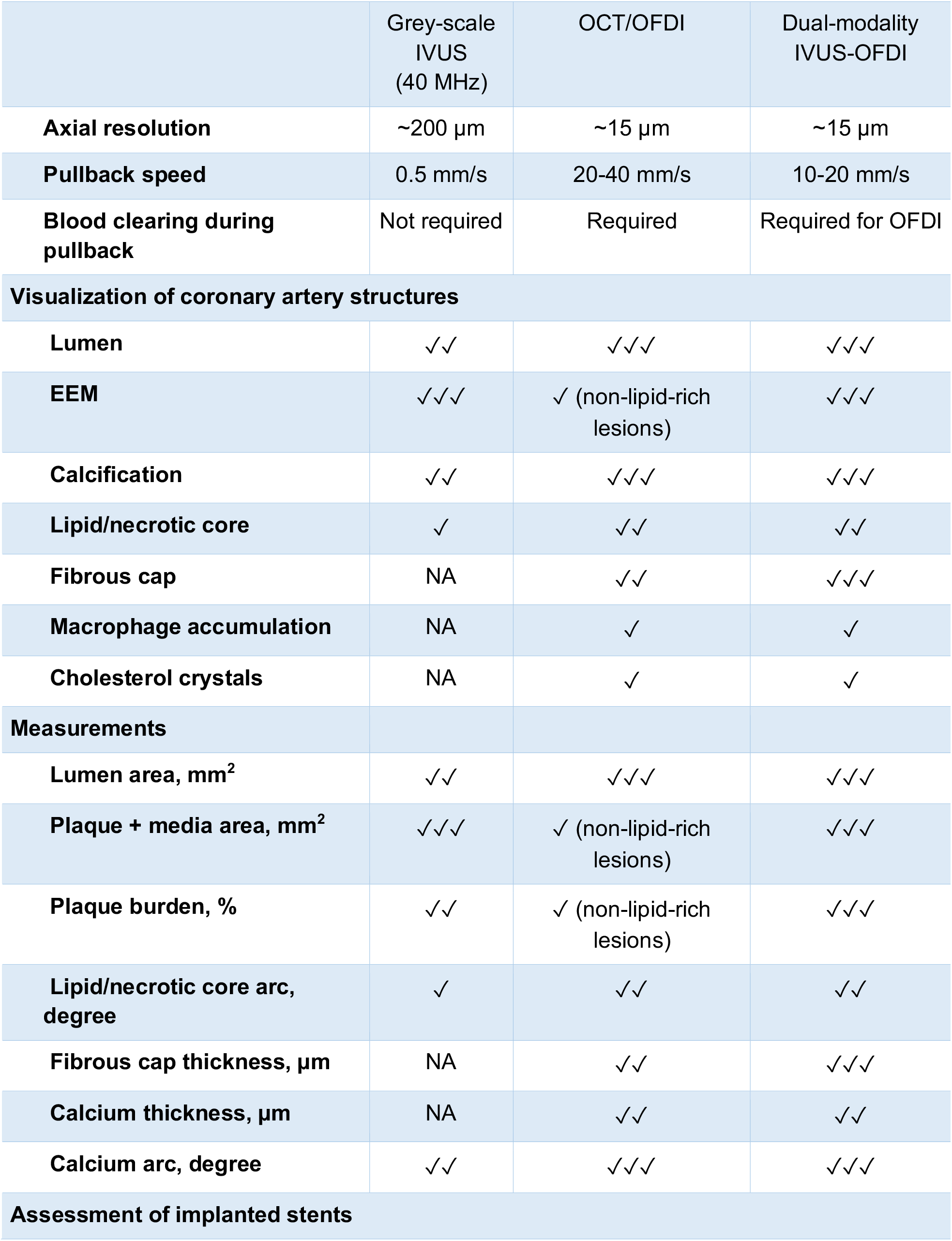
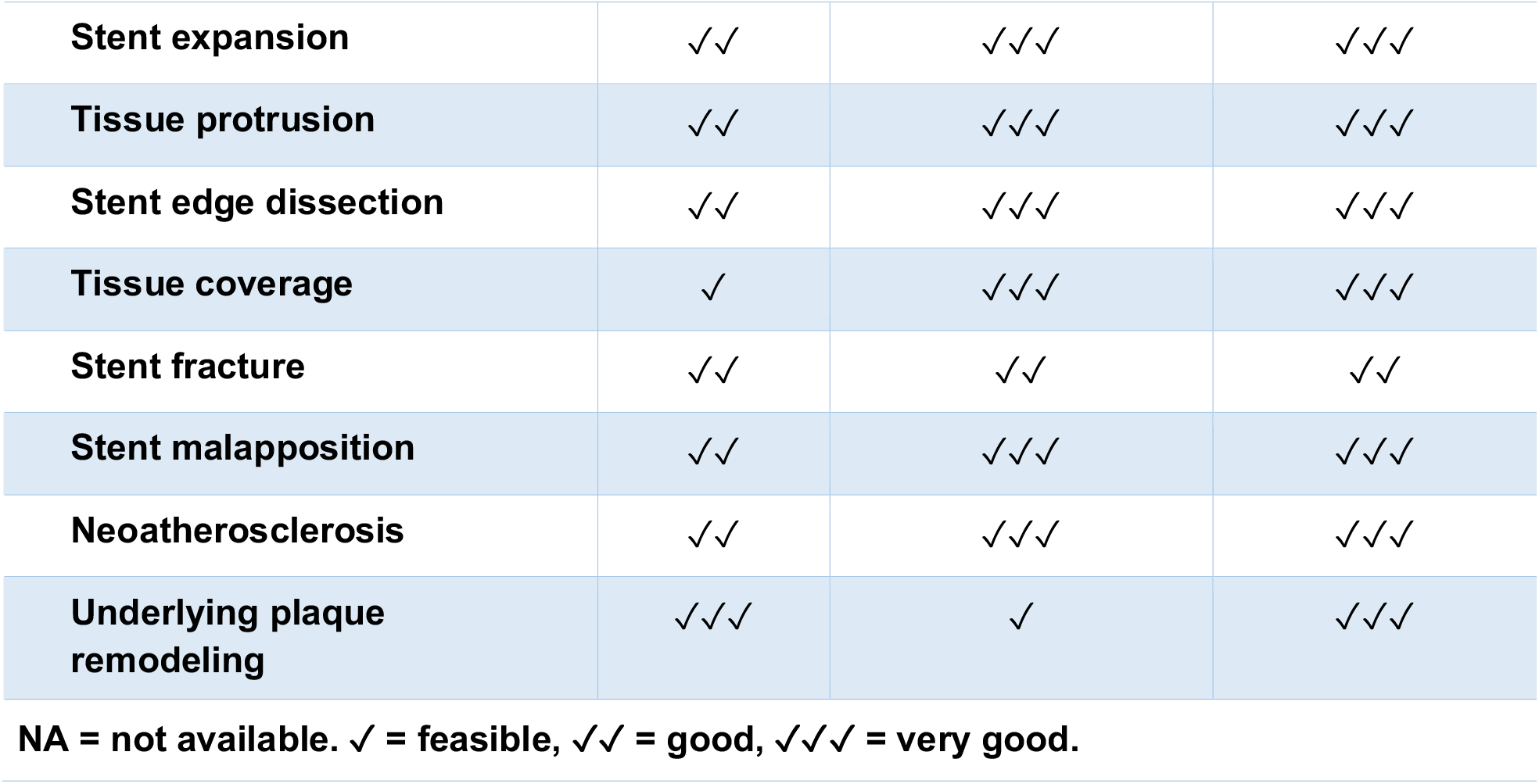
Comparison of IVUS, OFDI, and the dual-modality IVUS-OFDI in their ability to characterize coronary artery structures

|  | Grey-scale IVUS (40 MHz) | OCT/OFDI | Dual-modality IVUS-OFDI |
| --- | --- | --- | --- |
| **Axial resolution** | ~200 μm | ~15 μm | ~15 μm |
| **Pullback speed** | 0.5 mm/s | 20-40 mm/s | 10-20 mm/s |
| **Blood clearing during pullback** | Not required | Required | Required for OFDI |
| **Visualization of coronary artery structures** |  |  |  |
| **Lumen** | ✓✓ | ✓✓✓ | ✓✓✓ |
| **EEM** | ✓✓✓ | ✓ (non-lipid-rich lesions) | ✓✓✓ |
| **Calcification** | ✓✓ | ✓✓✓ | ✓✓✓ |
| **Lipid/necrotic core** | ✓ | ✓✓ | ✓✓ |
| **Fibrous cap** | NA | ✓✓ | ✓✓✓ |
| **Macrophage accumulation** | NA | ✓ | ✓ |
| **Cholesterol crystals** | NA | ✓ | ✓ |
| **Measurements** |  |  |  |
| **Lumen area, mm<sup>2</sup>** | ✓✓ | ✓✓✓ | ✓✓✓ |
| **Plaque + media area, mm<sup>2</sup>** | ✓✓✓ | ✓ (non-lipid-rich lesions) | ✓✓✓ |
| **Plaque burden, %** | ✓✓ | ✓ (non-lipid-rich lesions) | ✓✓✓ |
| **Lipid/necrotic core arc, degree** | ✓ | ✓✓ | ✓✓ |
| **Fibrous cap thickness, μm** | NA | ✓✓ | ✓✓✓ |
| **Calcium thickness, μm** | NA | ✓✓ | ✓✓ |
| **Calcium arc, degree** | ✓✓ | ✓✓✓ | ✓✓✓ |
| **Assessment of implanted stents** |  |  |  |
| **Stent expansion** | ✓✓ | ✓✓✓ | ✓✓✓ |
| **Tissue protrusion** | ✓✓ | ✓✓✓ | ✓✓✓ |
| **Stent edge dissection** | ✓✓ | ✓✓✓ | ✓✓✓ |
| **Tissue coverage** | ✓ | ✓✓✓ | ✓✓✓ |
| **Stent fracture** | ✓✓ | ✓✓ | ✓✓ |
| **Stent malapposition** | ✓✓ | ✓✓✓ | ✓✓✓ |
| **Neoatherosclerosis** | ✓✓ | ✓✓✓ | ✓✓✓ |
| **Underlying plaque remodeling** | ✓✓✓ | ✓ | ✓✓✓ |

NA = not available. ✓ = feasible, ✓✓ = good, ✓✓✓ = very good.

